# Transcranial direct current stimulation over the left inferior frontal gyrus improves sentence comprehension

**DOI:** 10.1101/2020.09.08.20190744

**Authors:** Eleni Peristeri, Zeyi Wang, Olivia Herrmann, Brian Caffo, Constantine Frangakis, Kyrana Tsapkini

## Abstract

**Background:** The left inferior frontal gyrus (IFG) has been shown to be involved in sentence comprehension in many studies through its involvement in both semantic and syntactic computations. However, causal evidence for its involvement in sentence comprehension is scarce. We used transcranial direct current stimulation (tDCS) to test the causal involvement of the left IFG in sentence comprehension in a group of individuals with primary progressive aphasia (PPA). These individuals participated in a tDCS study targeting lexical retrieval only, not sentence comprehension, therefore in the present study we report on far-transfer effects of tDCS in sentence comprehension.

**Objective:** We sought to determine whether tDCS over the left inferior frontal gyrus (IFG) coupled with lexical retrieval treatment may improve sentence comprehension in PPA.

**Method:** Within a sham-controlled, double-blind design, we tested whether 15 daily sessions of anodal tDCS over the left IFG may improve sentence comprehension in 27 people with PPA, and whether the tDCS effects were sustained up to two months post-treatment.

**Results:** We found that immediately post-treatment, and up to 2-months post-treatment, there was significantly larger improvement of sentence comprehension in the tDCS condition compared to sham. There were, however, differential effects of tDCS in each PPA variant and sentence-type. Importantly, participants with the epicenter of atrophy over the stimulated area (non-fluent PPA) benefited most from tDCS.

**Conclusion:** TDCS over the left IFG induces far-transfer effects and may improve sentence comprehension in PPA. We provide causal evidence that left IFG is a critical area for sentence comprehension.

## 1. Introduction

Successful development of effective treatments targeting language deficits in individuals with aphasia, especially in progressive disorders, depends on two elements: how long training effects are sustained and how much of what is learned generalizes (transfers) to other items, tasks, or functions [1]. There are generally two ways in which generalization is measured: improvement in items of the trained task not practiced during treatment (untrained items, near-transfer effects) and improvement in tasks that are not practiced (untrained tasks, far-transfer effects). In the latter, the untrained tasks may be related to the training task(s), especially if they depend on the same language or cognitive function [2]. For example, improvement in lexical retrieval could potentially induce improvement in both picture naming and noun-verb word generation, since they depend on the same computation of the left inferior frontal gyrus (IFG), i.e., strategic word selection and retrieval, as seminal neuroimaging studies by Thompson-Schill and colleagues and Petrides and colleagues had shown early on [3,4].

Far-transfer effects are particularly important in neurodegenerative conditions since time has a detrimental impact on several language and cognitive functions. Most treatment studies in primary progressive aphasia (PPA) have shown near-transfer effects in untrained items [5]. There have been only four behavioral treatment studies (not involving electrical stimulation) that investigated far-transfer effects in neurodegenerative conditions, such as PPA, and have shown limited effects [6–9]

Transcranial direct current stimulation (tDCS), conversely, has shown promising far-transfer effects, specifically in post-stroke aphasia [10,11]. In stroke populations, due to the disruption of normal functional and structural connections, tDCS has been successfully applied to compensatory areas [12] or over large brain areas [13]. Although still in its advent for neurodegenerative conditions, there is promise that tDCS may augment language therapy effects in PPA, and most tDCS studies have shown near-transfer effects, i.e., generalization to untrained items [14–19]. There is also evidence that tDCS effects over the left IFG generalize to untrained but related language tasks in PPA, such as generalization of a naming intervention to an untrained verbal fluency task [20,21]; we reasoned that this occurred because both tasks involve strategic retrieval and selection of the correct word amongst alternatives. However, far-transfer tDCS effects to untrained language functions and not only tasks, are reported in two small studies that showed a generic overall improvement. Wang and colleagues’ single case-study [22] was the first to report that anodal tDCS over the left IFG improved oral word comprehension, reading, word repetition and picture naming; however, targeted speech-language training was not combined with tDCS treatment. Gervits and colleagues [20] implemented tDCS over the left frontal area paired with a story narration task in 4 participants and found improvements in picture naming, sentence repetition and even grammatical comprehension. Given that the trained task (story narration), corroborates all above language functions, and the small sample size, it cannot be assumed that the improved tasks where not actually trained.

Since time is costly for both the brain and its function, in the present study we seek to determine whether tDCS may efficiently maximize treatment outcomes by inducing far-transfer generalization effects to untrained language functions. Our primary question was whether tDCS over the left IFG paired with a written naming/spelling task would induce far-transfer generalization effects in an untrained language function involving syntax and sentence comprehension. Our secondary question was whether the generalization effects would be different amongst PPA variants.

*Involvement of the left IFG in sentence comprehension: syntactic and semantic contributions*

Lesion, treatment and imaging studies provide evidence for the critical role of the left IFG in sentence comprehension. It has been shown that the left IFG is critical in sentence comprehension due to its role in both syntactic computations *and* accessing semantic information from temporal cortices [23,24]. The left IFG comprises areas important for both semantics (Brodmann’s area [BA] 45/47) and syntax (BA 44). Areas BA 45 and BA 47 have been shown to be involved in the processing of semantic information in functional magnetic resonance imaging(fMRI) studies by strategic access and selection of such information from the temporal lobes [3,4]. Furthermore, DTI studies have shown that this is possible through the extreme capsule fasciculus that runs from BA 45/47 to the mid portion of the superior temporal gyrus (STG)/middle temporal gyrus (MTG) [23,25,26]. Combined fMRI and diffusion tensor imaging (DTI) studies have indicated that this ventral route through the extreme capsule is most relevant for semantic aspects of sentence comprehension [24,27,28] such as the argument structure of verbs in a sentence (who did what to whom). For the syntactic aspects of sentence comprehension, fMRI and DTI studies highlight the role of the dorsal route for syntactic processing [24,29] in healthy controls and individuals with PPA [30]. Taken together, the left IFG (comprised of areas BA 44 and BA 45/47) seems to be in the ideal position to coordinate semantic and syntactic processing for sentence comprehension, i.e., the alignment between thematic roles and hierarchical ordering of syntactic phrases in complex, non-canonical sentences, such as passives [29,31,32].

With regard to treatment, a recent single-case treatment study employing a syntactic intervention on a patient with non-fluent PPA showed improvement of his production and comprehension of non-canonical (passive and object cleft) sentences with upregulation of his frontal network [33]. Additionally, two tDCS studies in healthy controls have provided evidence for the causal involvement of the left IFG in syntactic computations [34,35]. These studies implemented a single session of tDCS over the left frontal operculum (left IFG in Giustolisi and colleague’s’ study [34] and left prefrontal cortex in Hussey and colleagues’ study [35]) paired with a sentence-picture matching task [34] or with an ambiguous garden-path sentence parsing task [35]. In both studies, the tDCS group performed more accurately than sham.

Given the strategic involvement of the left IFG in syntactic and semantic computations during sentence comprehension, and the fact that the mechanism of tDCS is to modulate the (original or compensatory) function of the areas on which it is applied [36,37], we hypothesized that tDCS over the left IFG would improve sentence comprehension, even if not trained.

## 2. Methods

### 2.1 Participants

In total, 27 individuals with PPA participated in this study (11 female). There were 11 participants diagnosed with nfvPPA, 11 with lvPPA, and 5 with svPPA based on clinical assessment, neuropsychological and language testing, and MRI by experienced clinicians in specialized centers. All were right-handed, native English speakers, 50–80 years old who provided informed consent to participate in the clinical trial (http://ClinicalTrials.gov Identifier: NCT02606422), in compliance with the Johns Hopkins Hospital Institutional Review Board. Tables 1A and 1B show scores of each variant group for sex, age, education, years post onset of symptoms, and overall Frontotemporal Dementia Clinical Dementia Rating score (FTD-CDR) as well as language severity measures (Language FTD-CDR) [38].

**Table 1A:** Means (standard deviations) of demographics grouped at baseline (n = 27). *Fisher’s exact test used; FTD-CDR, Frontotemporal Dementia Clinical Dementia Rating Scale sum of boxes [38]; F, female; M, male. L, logopenic; N, nonfluent; S semantic; H.S., High School. ** F(1, 21) statistic due to 4 dropouts in phase 2 sham group.

|  | tDCS first | sham first | F(1, 25) | p-value |
| --- | --- | --- | --- | --- |
| Sex | 4 F, 7 M | 7 F, 9 M | * | 0.100 |
| Variant | 4 L, 4 N, 3 S | 7 L, 7 N, 2 S | * | 0.766 |
| Age at start of therapy (years) | 65.45 (8.80) | 69.88 (5.75) | 2.506 | 0.126 |
| Symptom onset before therapy (years) | 5.45 (3.69) | 4.56 (2.34) | 0.593 | 0.448 |
| Language Clinical Dementia Rating | 1.91 (0.92) | 1.81 (0.66) | 0.102 | 0.752 |
| Total severity (FTD-CDR) | 6.09 (3.26) | 7.16 (3.84) | 0.564 | 0.460 |
| Sessions in phase 1 | 12.73 (1.62) | 11.31 (1.49) | 5.471 | 0.028 |
| Sessions in phase 2 | 11.29 (1.89) | 11.06 (1.39) | 0.101** | 0.754 |

**Table 1B:** Means (standard deviations) of demographics grouped by PPA variant (n = 27). *Fisher’s exact test used. FTD-CDR, Frontotemporal Dementia Clinical Rating Scale sum of boxes [38]; F, female; M, male; s, sham; t, tDCS. ** F(2, 20) statistic due to 4 dropouts in phase 2 sham group (2 lvPPA, 2 nvPPA).

|  | nvPPA | lvPPA | svPPA | F(2, 24) | p-value |
| --- | --- | --- | --- | --- | --- |
| Sex | 4 F, 7 M | 5 F, 6 M | 2 F, 3 M | * | 1.000 |
| First-period condition | 4 tDCS | 4 tDCS | 3 tDCS | * | 0.766 |
|  | 7 Sham | 7 Sham | 2 Sham |  |  |
| Sessions in Phase 1 | 11.27 (1.42) | 12.09 (1.76) | 12.80 (1.79) | 1.648 | 0.213 |
| Years post-onset | 4.86 (2.71) | 4.23 (3.23) | 6.60 (2.58) | 1.140 | 0.336 |
| Age at Start of Therapy | 69.45 (6.52) | 66.18 (8.66) | 69.20 (6.26) | 0.601 | 0.556 |
| Language Severity (FTD-CDR) | 2.18 (0.60) | 1.36 (0.78) | 2.20 (0.45) | 5.067 | 0.015 |
| Total Severity (FTD-CDR) | 8.77 (3.86) | 4.86 (3.02) | 6.30 (1.60) | 4.086 | 0.030 |

### 2.2 Overall Design

The trial followed was a randomized, sham-controlled, crossover and double-blind design with two experimental conditions defined by stimulation type: language therapy plus anodal tDCS over the left IFG (tDCS condition) or language therapy plus sham tDCS (sham condition). Randomization was completed at baseline before treatment. We randomized stimulation condition within each variant every 4 participants. Each period of stimulation consisted of 12–15 consecutive daily weekday sessions, depending on patient’s availability and other life/health circumstances. An interval of 2 months separated the two treatment periods.

In the present report, we used only the first period of stimulation before the crossover because we had found some possible carryover effects in the main trial [39]. Participants were evaluated immediately before, immediately after, 2-weeks and 2 months post-stimulation. The 2-week follow-up evaluation was conducted online for some participants and missed for others, resulting in significantly fewer datapoints. Therefore, this evaluation point was omitted in the present analyses. Apart from the trained and untrained word sets for oral and written naming and spelling, we conducted a comprehensive neuropsychological and neurolinguistic evaluation (including sentence comprehension reported here) at each follow-up timepoint.

### 2.3 tDCS methods

We used a conventional tDCS approach to target the left IFG. The anode was placed over the left IFG and the cathode over the right cheek (extracephalic). Non-metallic, conductive electrodes, in saline-soaked sponges (5 cm x 5 cm), were held against the scalp by a pair of large, adjustable head straps and adhesive bandage tape (see montage in previous trial reports [39]). The anode was centered at F7 in line with the EEG 10–20 international system of electrode placement [40]. During the active tDCS condition, tDCS was applied to the scalp over the target using a battery-powered Soterix CT 1X1 tDCS device delivering 2 mA per minute (estimated current density 0.08 mA/cm^2^; estimated total charge 0.096 C/cm^2^). For the active tDCS condition, the stimulator was programmed to deliver current for 20 minutes for a daily maximum of 40 mA. For sham condition, sponge electrodes were applied in the same manner and current was ramped up to 2 mA and then 30 seconds later it was ramped down to 0 mA. Each daily treatment session lasted approximately one hour. Stimulation was delivered at the start of treatment, in conjunction with language therapy, and lasted 20 minutes; language therapy continued for an additional 25 minutes (45 minutes total). Participants were asked to rate their average pain intensity twice during each session (sham or tDCS) with the Wong-Baker FACES Pain Rating Scale.

### 2.4 Language Intervention

Language therapy involved an oral and written naming paradigm [41] adapted to emphasize spelling in a spell-study-spell procedure [42]. During a session, the participant was shown a picture on the computer, asked to name it orally, and then write the name. Semantic feature analysis [43], verbal repetition and spell-study-spell procedures [42] were implemented if the participant struggled to produce the target word in either modality. Individualized word sets created for each participant while maintaining the same procedures to address differing deficits across participants. For more details see previous reports of the trial’s main outcomes [39].

### 2.5 Sentence comprehension assessment

At each evaluation point (baseline, immediately after treatment, 2 weeks and 2-months post-treatment) participants were assessed using an extensive battery of standardized language and cognitive tasks.

As stated above, the treatment targeted lexical retrieval at the word level. The sentence comprehension task we report here, The SOAP (A Test of Syntactic Complexity), is a standardized task by Love and Oster [44]. It was, thus, not targeted in therapy (untrained) and presented only at each evaluation point.

Briefly, the SOAP requires individuals to listen to a sentence produced by the experimenter and point to the picture, amongst three options, that accurately corresponds to the meaning of the sentence. Options for pictures include the ‘matched’ (i.e., correct) picture, as well as two ‘mismatched’ pictures. The first ‘mismatched’ picture depicts the same action as the target sentence, but the thematic roles of the agent and patient are reversed. The second ‘mismatched’ picture includes characters or actions semantically unrelated to the target sentence. To avoid non-syntactic cues, all modifiers of characters in the sentences (e.g., hair color; see ‘brown hair’ in examples below) are repeated for each sentence and character in each sentence subset. Sentences are reversible, with animate agents in noun phrases (NPs), and matched for number of words to avoid differentiated working memory demands between syntactic conditions.

The task consists of 40 total sentences, with 10 trial sentences for each sentence-type condition: active sentences, passive sentences, subject relative sentences and object relative sentences (see examples 1–4 below).
1. The young boy with the brown hair grabs the man (Active)
2. The boy with the brown hair is grabbed by the man (Passive)
3. The man that grabs the little boy has brown hair (Subject-relative)
4. The man that the young boy grabs has brown hair (Object-relative)

Outcome measures in the analyses were the total accuracy score collapsing over sentence-types, and accuracy scores for each sentence-type.

### 2.6 Statistical Analyses

#### 2.6.1 Estimation of tDCS effects

Only data from the first period of stimulation were used to avoid impact of possible carryover effects or treatment-phase interaction due to possibly longer than expected lasting tDCS effects. The outcome measure (target of inference) was the average treatment effect (ATE), i.e. the additional, individual-specific tDCS effect over sham, δ_(T vs S)_ = E[Y|T = 1] - E[Y|T = 0] after taking into account the following covariates, where Y is the change in sentence comprehension scores from baseline and T is the treatment assignment indicator (valued as 1 for tDCS, 0 for sham).

The following *covariates* were used to improve the efficiency of tests: baseline sentence comprehension (shown by previous research to influence tDCS effects in PPA [20,45]), PPA variant, number of treatment sessions, sex, age, years post onset of symptoms, and total FTD-CDR severity and language severity measures.

We applied the Targeted Maximum Likelihood Estimation (TMLE) [46] for the estimation and inference of ATE for each participant using the TMLE R package [47]. Compared to other available consistent estimation strategies for such setting, our method choice was in order to improve the inference efficiency by accounting for baseline covariate information with flexible models while reducing subjective model selection. To derive the final estimation, an outcome model and a propensity score model were fitted separately with cross-validation. The candidate models included a regression with all the main terms and the stepwise regressions with forward and backward directions, and the selection was based on cross-validation prediction accuracy. The estimations, standard errors, Z-test statistics, p-values, and 95% confidence intervals are reported (see Table 2).

At two months post, three dropouts were assumed as Missing at Random (MAR).

**Table 2.** Results of variant subgroup analysis for immediately after and 2 months post-intervention.

| | Estimate | SE | $\chi^2(1)$ | *p* | Lower | Upper |
| --- | --- | --- | --- | --- | --- | --- |
| T vs S, nfvPPA, After | 6.04 | 2.23 | 7.31 | 0.007 | 1.66 | 10.41 |
| T vs S, lvPPA, After | 2.26 | 2.17 | 1.08 | 0.299 | -2.00 | 6.51 |
| T vs S, svPPA, After | 0.60 | 2.98 | 0.04 | 0.841 | -5.24 | 6.44 |
| T vs S, nfvPPA, 2month | 5.69 | 2.44 | 5.43 | 0.020 | 0.90 | 10.47 |
| T vs S, lvPPA, 2month | 1.90 | 2.36 | 0.65 | 0.421 | -2.73 | 6.54 |
| T vs S, svPPA, 2month | 0.25 | 2.90 | 0.01 | 0.932 | -5.44 | 5.94 |

#### 2.6.2 Variant and sentence-type effects

We further investigated how the tDCS effects on changes in sentence comprehension are modified by each variant subgroup and by each sentence-type at different time points. We assumed MAR for dropouts at two months post and applied the linear mixed model (LMM) with random subject intercepts for each of the four SOAP sentence-types. Each fixed effect model consisted of treatment, time point, treatment time interaction, treatment variant interaction, time variant interaction, and baseline covariates (see section 2.6.1). Additional tDCS effects were evaluated at different time points (immediately after and 2 months post) for each variant subgroup (nfvPPA, svPPA, lvPPA) and for each sentence-type (Subject‐relative, Object‐relative, Active, Passive). Estimations, standard errors, Wald test statistics, p-values and 95% confidence intervals are reported in Tables 4–7.

#### 2.6.3 Selection of baseline predictors for the Potentially Heterogeneous tDCS effects

For each individual, the individual-specific additional tDCS effect (i.e., the difference between the potential change in sentence comprehension if the participant was assigned to tDCS versus sham) may be heterogeneous. The heterogeneity of the tDCS effect on change in sentence comprehension scores was captured by the conditional average treatment effect (CATE), E[Y(T = 1) – Y(T = 0) | X], where Y(T = 1) and Y(T = 0) are the counterfactual changes depending on the treatment group (T = 1 for tDCS, T = 0 for sham), and X represents the predictors that may interact with the individual-specific tDCS effect. We tested whether any of the baseline factors (baseline SOAP score, PPA variant, number of treatment sessions, sex, age, years post onset of symptoms, and total FTD-CDR severity and language severity measures) were *predictors* of the individual tDCS effect.

To select the baseline factors that are predictive for CATE, a nonparametric regression method using jackknife pseudovalues was applied [48]. The candidate factors were selected depending on whether they were predictive to the pseudovalues, where fully observed pseudovalues act as an unbiased transformation of the unobservable individual tDCS effect, i.e. E[U_i_ | X = x_i_] = E[Y_i_(T = 1) – Y_i_(T = 0) | X = x_i_], where U_i_ = 2Y_i_T_i_ – 2Y_i_(1-T_i_) is the jackknife pseudovalue of the *i*-th subject, and x_i_ represents the observed factor values from the *i*-th subject. Linearity was assumed and variable selection was conducted based on the leave-one-out cross validated (LOOCV) predictive R-squared. At each step of the forward selection, a threshold of 0.1 on the R-squared increase was applied to stop the selection procedure, otherwise the variable with the largest R-squared increase was selected. The predictive R-squared and the root mean squared error (RMSE) of the current step, as well as the increase in predictive R-squared compared to the last step, are reported for each round of the variable selection.

## 3. Results

### 3.1 tDCS Tolerability

Participants reported some light tingling, itching, or discomfort from the stimulation, but no episodes of intolerability or adverse effects occurred. The maximum reported FACES pain rating for each daily session was averaged across sessions and participants, with a tDCS mean pain rating of 2.21 (standard deviation 2.48, range 0–10) and a sham mean rating of 2.14 (standard deviation 2.13, range 0–10).

### 3.2 Evaluation of tDCS effects in sentence comprehension immediately after and 2 months post-intervention

We present the results of participants’ performance in SOAP immediately after and 2 months post the intervention. Sentence comprehension scores ranged from 0–40, the sum of responses in the subject‐relative, object, relative, active, and passive sentences of the test (10 sentences per type, see section *2.5*). All statistical analyses were performed the changes from the baseline in the summed scores.

Restricting to the first-phase data, from before to immediately intervention, the additional effect of tDCS vs. sham on SOAP changes from baseline was significant (Estimate = 4.96, 95% CI [0.91, 9.01], SE = 2.07, *Z* = 2.40, *p* = 0.016) adjusted for all the covariates (baseline SOAP score, PPA variant, number of treatment sessions, sex, age, years post onset of symptoms, and total FTD-CDR severity and language severity measures) as listed in Section *2.1*. A marginally significant tDCS effect was still found for two months post-intervention (Estimate = 4.56, 95% CI [-0.07, 9.19], SE = 2.36, *Z* = 1.93, *p* = 0.053).

### 3.3 Variant effects

Participants with nfvPPA appeared to be the most responsive variant group, with the additional tDCS effect estimated as 6.03 at immediately after and 5.68 at 2 months post (see Table 2).

### 3.4 Sentence-type effects

Participants with nfvPPA appeared to be the most responsive variant group in passives, with the effect sizes estimated as 1.95 at immediately after and 2.05 at 2 months post (see Table 3, Figure 1).

**Table 3.** Results of passive sentences per variant subgroup analysis for immediately after and 2 months post the intervention.

| | Estimate | SE | $\chi^2(1)$ | *p* | Lower | Upper |
| --- | --- | --- | --- | --- | --- | --- |
| T vs S, nfvPPA, After | 1.95 | 0.97 | 4.01 | 0.045 | 0.04 | 3.86 |
| T vs S, lvPPA, After | 0.35 | 0.93 | 0.14 | 0.709 | -1.48 | 2.17 |
| T vs S, svPPA, After | 0.12 | 1.28 | 0.01 | 0.928 | -2.40 | 2.63 |
| T vs S, nfvPPA, 2month | 2.05 | 1.06 | 3.77 | 0.052 | -0.02 | 4.13 |
| T vs S, lvPPA, 2month | 0.45 | 1.02 | 0.20 | 0.656 | -1.54 | 2.44 |
| T vs S, svPPA, 2month | 0.22 | 1.24 | 0.03 | 0.860 | -2.22 | 2.66 |

**Figure 1.**
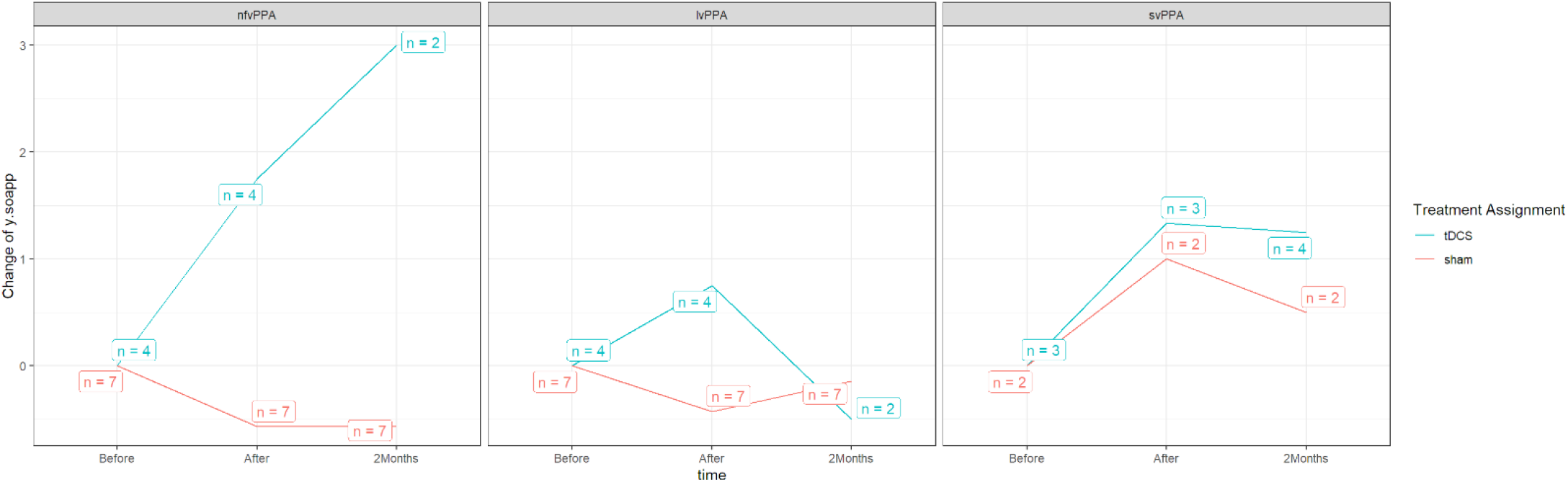
Trend plot of sentence comprehension change per variant in passives (shown on the ordinate, y-axis) from baseline (Before) for tDCS vs. sham, immediately after intervention (After), and 2 months post intervention (2Months) shown in the abscissa (x-axis) for each PPA variant group; the statistical sample (number of participants) is indicated in boxes for both sham and tDCS.

Tables 4–6 present effect sizes across groups for actives, object relatives, and subject relatives immediately after tDCS and at 2 months post-treatment.

**Table 4.** Results of active sentences per variant subgroup analysis for immediately after and 2 months post the intervention. The first phase data was used.

| | Estimate | SE | $\chi^2(1)$ | *P* | Lower | Upper |
| --- | --- | --- | --- | --- | --- | --- |
| T vs S, nfVPPA, After | 1.68 | 0.75 | 4.93 | 0.026 | 0.20 | 3.15 |
| T vs S, lvPPA, After | 0.43 | 0.76 | 0.32 | 0.572 | -1.05 | 1.91 |
| T vs S, svPPA, After | -0.21 | 0.98 | 0.04 | 0.834 | -2.13 | 1.72 |
| T vs S, nfVPPA, 2month | 1.59 | 0.85 | 3.52 | 0.061 | -0.07 | 3.26 |
| T vs S, lvPPA, 2month | 0.35 | 0.85 | 0.17 | 0.684 | -1.32 | 2.02 |
| T vs S, svPPA, 2month | -0.29 | 0.95 | 0.09 | 0.764 | -2.16 | 1.58 |

**Table 5.** Results of object relative sentences per variant subgroup analysis for immediately after and 2 months post the intervention. The first phase data was used.

| | Estimate | SE | $\chi^2(1)$ | *p* | Lower | Upper |
| --- | --- | --- | --- | --- | --- | --- |
| T vs S, nfVPPA, After | 1.31 | 0.99 | 1.74 | 0.19 | -0.64 | 3.26 |
| T vs S, lvPPA, After | 1.57 | 0.99 | 2.50 | 0.11 | -0.38 | 3.52 |
| T vs S, svPPA, After | 0.12 | 1.29 | 0.01 | 0.93 | -2.40 | 2.64 |
| T vs S, nfVPPA, 2month | 0.92 | 1.09 | 0.71 | 0.40 | -1.22 | 3.07 |
| T vs S, lvPPA, 2month | 1.18 | 1.09 | 1.19 | 0.28 | -0.95 | 3.32 |
| T vs S, svPPA, 2month | -0.27 | 1.25 | 0.05 | 0.83 | -2.72 | 2.19 |

**Table 6.**
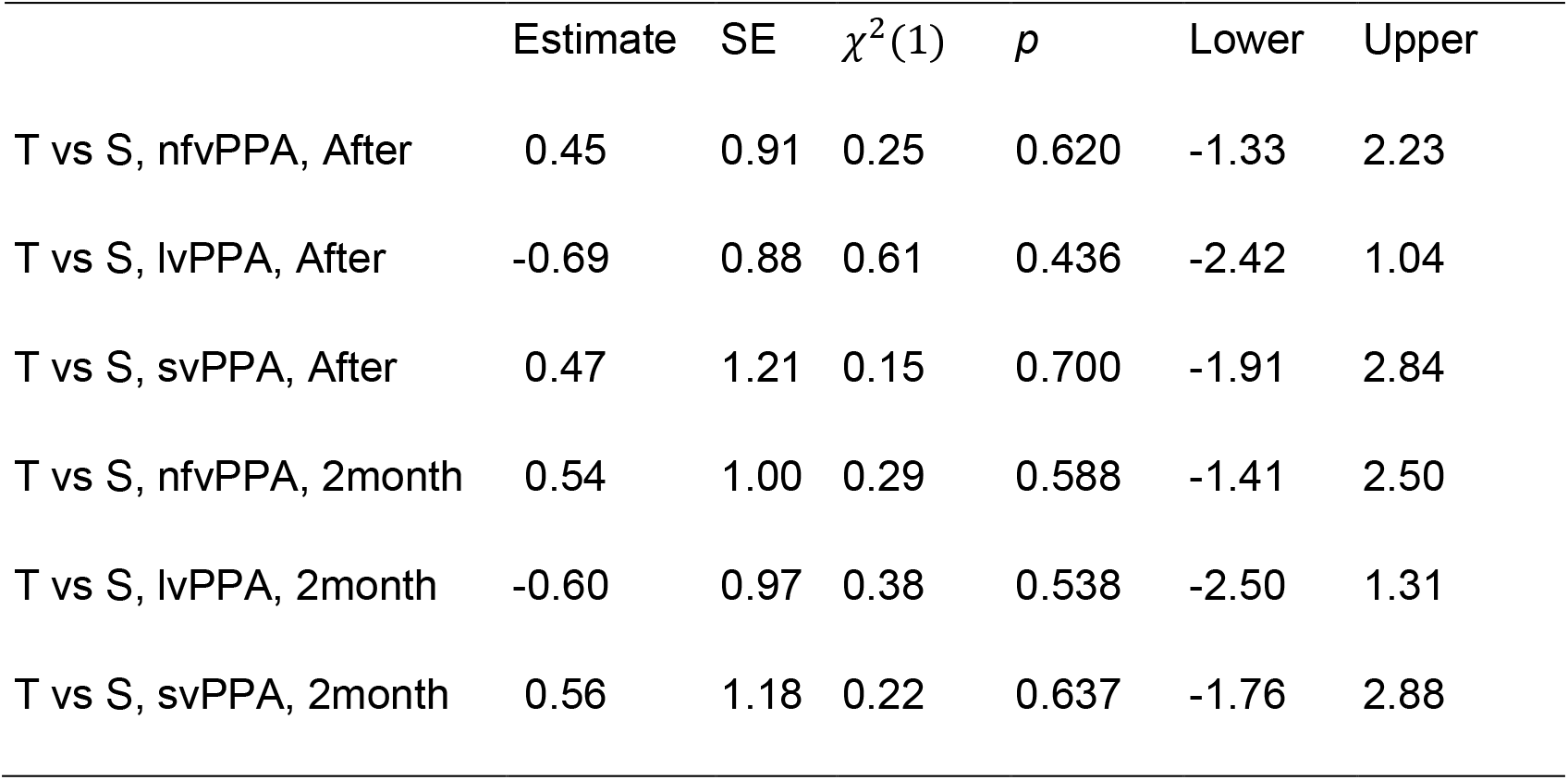
Results of subject relative sentences per variant subgroup analysis for immediately after and 2 months post the intervention.

### 3.5 Prediction of Potentially Heterogeneous tDCS effects: baseline factors

Two baseline factors were selected as predictors for the individual tDCS effect, the indicator of being svPPA and years post-onset (predictive R-squared increases: 0.129 and 0.141, respectively, see Table 7). This hints at potential predictiveness of a patient’s severity, which has been found associated with years post-onset in previous research [20,45].

**Table 7.**
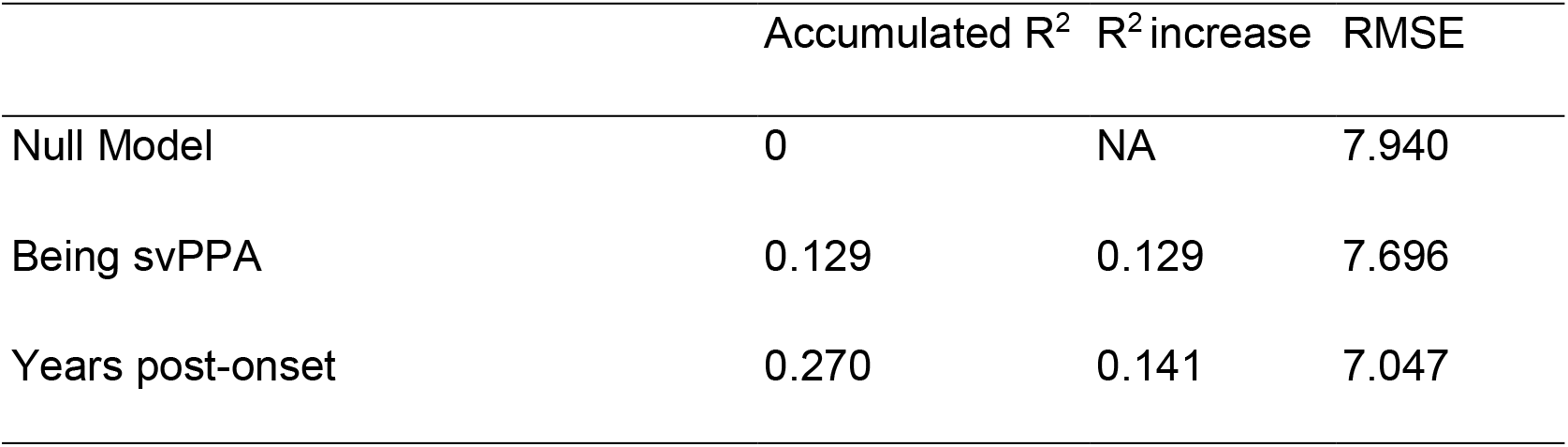
Baseline factors for individual tDCS effect prediction.

## 4. Discussion

In this study, we addressed the question of whether tDCS effects may generalize to far-transfer language functions. We sought to determine whether the additional effects of tDCS over the left IFG coupled with a lexical retrieval therapy (targeting written naming/spelling) may improve performance in an untrained sentence comprehension task in individuals with PPA. The main finding is that stimulation over the left IFG induced far-transfer effects in sentence comprehension that sustained for up to 2 months. Furthermore, the effect was most pronounced in participants with nfvPPA, whose epicenter of atrophy is over the left IFG, and specifically in passive sentences. The far-transfer effect is discussed with regard to the role of the left IFG in sentence comprehension.

The present study aligns with previous claims of far-transfer tDCS effects in PPA to the largest to-date group of patients. In particular, Wang and colleagues [22] found that anodal tDCS over the left IFG resulted in improved oral word comprehension, reading, word repetition and picture naming in a single patient with nfvPPA. Moreover, Gervits and colleagues [20] studied the effects of tDCS with the anode placed over the left frontotemporal region in six participants with PPA (2 with nfvPPA, 4 with lvPPA). Stimulation resulted in improvements in semantic processing and grammatical comprehension, some of which were maintained up to 12 weeks post-stimulation. Studies showing generalization effects of tDCS in untrained items (also called near-transfer effects) are considerably more [14,16,39] and show greater and longer-lasting improvements in naming performance for individuals with PPA compared to sham condition. To the best of our knowledge, our study is the first to report improvement in sentence comprehension (untrained task) lasting up to 2-months post-stimulation over the left IFG compared to a sham control.

Our findings of tDCS-driven improvement in sentence comprehension also align with previous tDCS studies with healthy individuals that found significant improvements in comprehension of sentences of higher syntactic complexity after tDCS over the left IFG paired with a sentence comprehension task [34,35]. The present study extends these findings to the largest to-date group of PPA individuals who did not receive targeted sentence comprehension treatment. Importantly, we provide causal evidence on the involvement of the left IFG in higher-level language comprehension since the tDCS effects were independent from the treatment (lexical retrieval).

As summarized in the introduction, several lesion, fMRI and DTI studies have provided evidence that left IFG plays a pivotal role in sentence comprehension due to its position that comprises both areas of semantic selection (BA 45/47) through the ventral language route, as well as areas of hierarchical ordering of syntactic phrases (BA 44) through the dorsal language route [27,29,31,32,49]. Although the evidence in PPA for sentence processing is more in favor of the dorsal route [30,50], probably due to the type of sentences studied, the involvement of the left frontal operculum is indisputable [30,33]. The present study provides strong evidence not only on the role of the left IFG in sentence comprehension in PPA but also on the causal involvement of the left IFG (with its semantic and syntactic computations) in sentence comprehension and its rehabilitation in a neurodegenerative condition.

Interestingly, improvement in sentence comprehension was more pronounced in passives. Passives include a single verb (a single thematic role assigner) in contrast to object relatives, which include two verbs and, thus, two syntactic dependencies to be established between moved NPs and their respective gaps. The lack of improvement in non-canonical object relatives for the PPA participants but not in passives may be explained by possible processing bottlenecks driven by the high short-term memory (STM) demands posed by 2-verb thematic role assignments needed for comprehension of sentences with object relatives. This interpretation aligns with previous findings showing that deficits in non-canonical structures with object and subject relatives are due to deficits in verbal STM abilities present in both nfvPPA and lvPPA [51]. In line with this interpretation, the neural substrate of verbal STM is not in the left IFG but rather in the supramarginal gyrus [52–54], and we did not stimulate this area. Therefore, it seems that improvement was specific to the computation of the stimulated area only.

Besides the fact that tDCS over the left IFG was related to improved accuracy mostly in passives, there were also differential effects of tDCS across variants: nfvPPA with more atrophy over frontal areas showed larger improvement [53]. While counterintuitive at first glance, there is accumulating evidence from ours and other groups that local atrophy does not correlate with either current distribution [55], functional connectivity [56,57], or even tDCS effects [20,39]. Therefore, tDCS may be more effective for individuals with vast tissue integrity loss, or over such compromised brain areas, augmenting their lower baseline, as proposed earlier [20]. Another possible explanation is that the effectiveness of tDCS relies on compensatory brain areas that remain intact. In general, individuals with nfvPPA have the least atrophied temporal regions [53], and these regions have been shown to be required for semantic aspects of sentence comprehension [27,32,58]. Whether the effects were caused by remote compensatory areas or the diffuse nature of the conventional tDCS effect, remains a question for further research.

Two limitations are noteworthy. First, we chose to stimulate the left IFG for all PPA variants given its key role in syntax. For this reason, we could not determine effects of delivering stimulation over other loci of atrophy in the individuals with PPA or in other areas of syntactic processing in the brain. Second, the electrode patches covered larger cortical areas than the left IFG, so there might have been current spread to adjacent non-targeted areas that might have affected the language rehabilitation process. Future studies should limit this possible effect of tDCS on adjacent areas with better spatial resolution protocols.

## 5 Conclusion

In 27 people with PPA, effects of tDCS over the left IFG transferred to an untrained sentence comprehension task. The present study offers novel evidence in favor of the causal involvement of the left IFG in syntactic computations during sentence comprehension and the sustainability of the language gains for individuals with neurodegenerative diseases. Further work investigating the precise mechanisms that might underlie these tDCS effects is needed.

## Data Availability

Data sharing agreement: We will make the data and associated documentation available to users only under a data-sharing agreement that provides for: (1) a commitment to using the data only for research purposes and not to identify any individual participant; (2) a commitment to securing the data using appropriate computer technology; (3) proper acknowledgment of data source; and (4) a commitment to destroying or returning the data after analyses are completed.

## Acknowledgements

We would like to thank our participants and referring physicians for their dedication and interest in our study. Funding: This work was supported by grants from the Science of Learning Institute at Johns Hopkins University and by the National Institutes of Health (National Institute of Deafness and Communication Disorders) through award R01 DC014475 to KT. The authors declare no Conflict of Interest.

